# Genetic Ancestry and Risk of Atrial Fibrillation in Individuals of Black Ethnicity in the UK Biobank

**DOI:** 10.1101/2025.07.12.25331418

**Authors:** Chang Liu, Yan V. Sun, Linzi Li, Lindsay J. Collin, Amit J. Shah, Alvaro Alonso

## Abstract

**Background:** Black individuals have a lower incidence of atrial fibrillation (AF) than White individuals, despite a higher burden of traditional risk factors. Prior studies have suggested that European genetic ancestry may contribute to this paradox, but findings have been inconsistent.

**Methods:** We examined the association between European genetic ancestry and incident AF among 6,920 UK Biobank (UKB) participants who self-identified as Black and were free of AF at baseline. European ancestry proportions were estimated by comparing participants to HapMap reference populations and were analyzed both continuously and categorically using Cox proportional hazards models. Non-linear associations were evaluated using flexible hazard ratio spline curves. We then conducted a meta-analysis combining these results with those from the Atherosclerosis Risk in Communities Study, the Cardiovascular Health Study, and the Women’s Health Initiative (WHI).

**Results:** During a median follow-up of 13.7 years, 205 (3%) participants developed incident AF. Each 10% increase in European ancestry was nominally associated with increased AF risk (HR 1.07, 95% CI: 0.95-1.20), and individuals in the highest ancestry category had a higher AF incidence rate. Random-effects meta-analysis of all four cohorts yielded a pooled relative risk (RR) of 1.09 (95% CI: 0.99-1.21), with substantial heterogeneity largely driven by the WHI study. Excluding WHI resulted in a pooled estimate (RR 1.14, 95% CI: 1.05-1.23) without heterogeneity.

**Conclusion:** Our findings suggest a modest association between European genetic ancestry and increased AF risk among admixed Black individuals. Future studies should confirm these results, explore underlying mechanisms, and assess their clinical implications.

## INTRODUCTION

Atrial fibrillation (AF) is a common arrhythmia and a well-established risk factor for stroke, heart failure, and other adverse outcomes.^1^ Its risk factors include older age, male sex, high blood pressure, obesity, diabetes and others.^2^ Epidemiologic studies in the United States have consistently shown that Black and Hispanic individuals have lower rates of AF compared with White individuals, despite having a higher burden of traditional AF risk factors.^3–6^ A recent analysis of the UK Biobank (UKB) found a similar pattern, with individuals of self-reported White race experiencing a higher incidence of AF compared with those of Black or Asian race groups.^7^

An analysis of the Atherosclerosis Risk in Communities (ARIC) Study and the Cardiovascular Health Study (CHS) found that among Black participants with genetic admixture, a higher proportion of European genetic ancestry was associated with an increased risk of AF, suggesting that ancestry-related genetic factors may play a role in this racial difference.^8^ However, a subsequent analysis of the Women’s Health Initiative (WHI) cohort did not replicate these findings.^9^ As a result, the relationship between genetic ancestry and AF risk remains unclear.

To further explore the potential role of genetic ancestry in explaining racial differences in AF risk, we examined the association between European genetic ancestry and AF incidence among UKB participants who self-identified as Black. Additionally, we conducted a meta-analysis incorporating the current results to those from the ARIC, CHS, and WHI cohorts, to provide a more comprehensive assessment of this relationship.

## METHODS

### The UK Biobank (UKB) cohort

The UKB is an ongoing, large-scale prospective cohort study based in the United Kingdom that enrolled over 500,000 participants aged 40-69 years between 2006 and 2010.^10^ The Ethics approval for UKB was provided by the North West Multi-centre Research Ethics Committee, and all participants provided written informed consent. The study collected extensive data through questionnaires, physical measurements, blood biomarker assays, imaging, genome-wide genotyping, and longitudinal follow-up.^10^ The cohort was linked to Hospital Episode Statistics (HES) data, primary care data and the death registry, which include information of death date and both primary and secondary causes.

This analysis was restricted to participants who self-identified as “Black or Black British”, “Caribbean”, “African”, or “Any other Black background” in response to the standard questionnaire on ethnic background at enrollment. Participants with genomics data, genetically unrelated (>3^rd^ degree relatedness), and without prevalent AF were included in this study.

### AF ascertainment

The UKB mapped first occurrence of disease diagnosis to International Classification of Diseases, 10th Revision (ICD-10) codes, integrating HES, primary care, death registry, and self-reported medical conditions. AF was defined through linkage with these records and by the presence of ICD-10 code I48, with the date of AF onset determined by its first occurrence in any linked dataset. Prevalent AF was defined as AF first recorded on or before enrollment, while incident AF was defined as AF first recorded after enrollment. Follow-up time was defined as time from enrollment to incident AF, lost to follow-up, death or the end of follow up as of September 2023.

### Determination of genetic ancestry

We performed ancestry estimation using ADMIXTURE,^11^ a model-based clustering method that estimates individual ancestry proportions from genome-wide genotype data. The analysis was conducted in supervised mode, using predefined ancestral reference populations to guide the estimation process. A total of 770 unrelated individuals from the HapMap phase 3 reference data were used as ancestry reference populations, including Utah residents with Northern and Western European ancestry from the CEPH collection (CEU); the Han Chinese in Beijing, China (CHB); the Chinese in Metropolitan Denver, Colorado (CHD); Japanese in Tokyo, Japan (JPT); Maasai in Kinyawa, Kenya (MKK); Luhya in Webuye, Kenya (LWK); and Yoruba in Ibadan, Nigeria (YRI). To reduce the number of highly correlated variants, we performed pruning of single nucleotide polymorphisms (SNPs) based on linkage disequilibrium (LD) using PLINK.^12^ Starting from all genetic variants with minor allele frequency > 5%, a sliding window of 50 consecutive SNPs was used, with the window shifted by 5 SNPs at each step, and SNP pairs with high LD (an r² value greater than 0.1) were pruned to reduce redundancy. Individuals self-reported as Black with a genetic ancestry proportion < 15% African ancestry (combination of MMK, LWK and YRI ancestry proportions) were excluded from the analysis (n = 33). A total of 73,693 independent genetic variants were included in this analysis. Additionally, a principal component analysis was performed using the pruned genomics data to visualize the population structure comparing the reference populations and the UKB participants.

### Covariates

Age, sex, education, total household income before tax, current smoking, blood-pressure lowering medication use were self-reported at enrollment. Height, weight, systolic, and diastolic blood pressure were measured at the baseline visit. Body mass index (BMI) was calculated as weight in kilograms divided by height in meters squared. Education was categorized as high (college or university degree; NVQ, HND, or HNC or equivalent, other professional qualifications [e.g., nursing, teaching]), low (A levels / AS levels or equivalent; O levels / GCSEs or equivalent; CSEs or equivalent; none of the above) and missingness was treated as a separate category. Household income was categorized as high (≥ £31,000), low (≤ £ 30,999), and missing. Baseline prevalence of type 2 diabetes (ICD-10 E11), coronary artery disease (ICD-10 I20 - I25), and heart failure (ICD-10 I50) were determined by ICD-10 codes from linkage to HES, primary care, death registry, self-reported medical conditions, and comparing the earliest date of onset to the enrollment date.

### Statistical analysis

Baseline characteristics of the study population were summarized by levels of European genetic ancestry and incident AF status using means and standard deviations for continuous variables and counts with percentages for categorical variables. Differences across groups were compared using one-way ANOVA, two-sample t test and Wilcoxon rank-sum test for continuous variables and chi-squared tests for categorical variables where appropriate.

We evaluated the association of European genetic ancestry with the incidence of AF using Cox proportional hazards model among eligible participants self-reporting Black ethnicity. European genetic ancestry was modeled as a continuous variable (per 10% higher). Model 1 adjusted for age at enrollment, sex (male vs. female), education (high vs. low vs. missing), income (high vs. low vs. missing). Model 2 additionally adjusted for current smoking, BMI, height, systolic blood pressure, diastolic blood pressure, blood pressure lowering medication use, prevalent type 2 diabetes, prevalent coronary artery disease, and prevalent heart failure. We conducted additional analyses categorizing European genetic ancestry into three groups using cutoffs of 11.26% (67th percentile) and 50%, including low (0.001%-11.25%), medium (11.26%-49.80%) and high (50.10%-73.43%[maximum]). The 11.26% cutoff was selected based on the overall distribution of European genetic ancestry and its non-linear association with incident AF risk, with a notable shift in risk observed around this threshold. The 50% cutoff was used to define individuals with predominant European ancestry, which is commonly applied in large-scale genetic and admixture studies to stratify populations based on majority ancestry. The groups of medium and high European genetic ancestry were compared with the low European genetic ancestry, using the same covariate adjustment used in Models 1 and 2. Sex-stratified analysis and analysis among only postmenopausal women were also performed.

We explored dose-response curves modeling European genetic ancestry as a spline term using the software smoothHR^13, 14^, based on the fully adjusted Model 2. This approach generated flexible hazard ratio curves that capture potential non-linear associations between ancestry and AF risk, allowing for the identification of inflection points along the exposure continuum.

We synthesized the results from the present analysis with those from the three previous studies reporting associations between European genetic ancestry and AF risk in Black individuals (ARIC, CHS, WHI).^15, 16^ Fully-adjusted risk ratios per 10% increase in European ancestry were pooled using the *metan* command in Stata.^17^ We first estimated a fixed-effect model with inverse of variance weighting and then a DerSimonian-Laird random-effects model because design heterogeneity made between-study heterogeneity plausible (ARIC, CHS and UKB only included incident cases, while WHI include prevalent and incident cases; WHI only included women). Heterogeneity was assessed using the I^2^ statistic and Cochran’s Q test.^18^ To assess the impact of each individual study on the overall summary estimate, we conducted a leave-one-out analysis by iteratively removing each study and recalculating the pooled risk ratio and heterogeneity statistics.

## RESULTS

The study cohort included 6,920 Black participants free of AF at baseline. The workflow of inclusion and exclusion is shown in **Supplementary Figure 1**. The mean age was 51.9 years (standard deviation [SD] 8.0), and 43.5% were male. The median follow-up time was 13.7 years (interquartile range [IQR] 13.2-14.5). During follow-up, a total of 205 individuals (3.0%) developed incident AF. European genetic ancestry proportions ranged from 0.001% to 73.43%, with a median of 6.3% (IQR 3.0%-15.0%). The overall distribution of European genetic ancestry is shown in **Figure 1**. Principal component analysis showed a clear alignment between UKB self-reported Black participants and the HapMap 3 reference populations of MKK, LWK, and YRI, **Supplementary Figure 2**.

**Figure 1.**
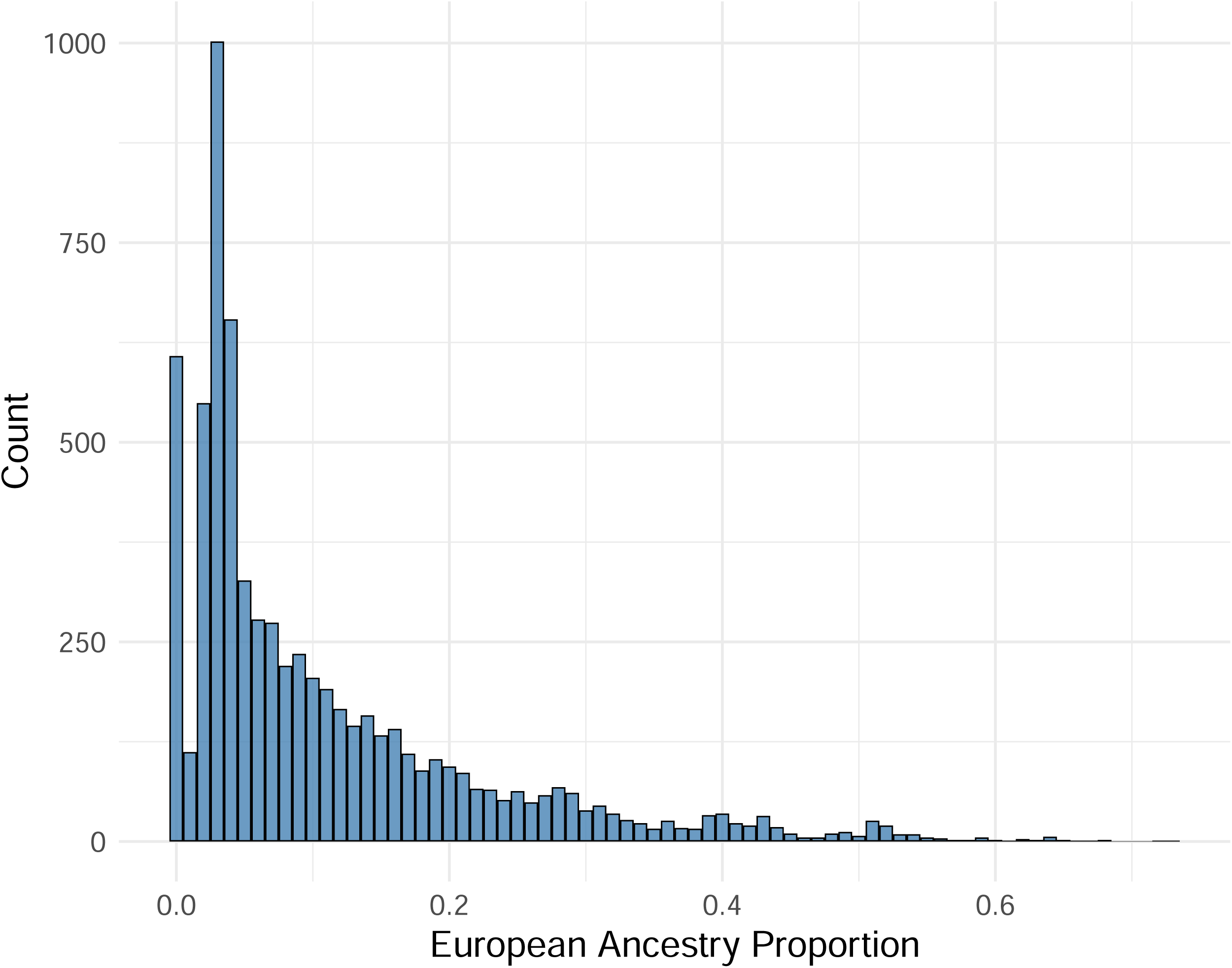
Distribution of European genetic ancestry among the UK Biobank participants self-reported as Black.

Baseline characteristics stratified by European ancestry categories are presented in **Table 1**, and by AF incidence status in **Table 2**. Participants with higher proportions of European ancestry were more likely to smoke, have lower BMI, lower blood pressure, and less likely to use blood pressure lowering medications. Incident AF cases were older, more likely to be male, less likely to smoker, taller, and had higher prevalence of cardiometabolic risk factors such as elevated BMI, systolic blood pressure, diabetes, and history of coronary artery disease and heart failure.

**Table 1.** Baseline characteristics by categories of European genetic ancestry proportions.

| Characteristics |  | Overall<br>N = 6,920 | Low*<br>N = 4,613 | Medium*<br>N = 2,198 | High*<br>N = 109 | P |
| --- | --- | --- | --- | --- | --- | --- |
| European genetic ancestry proportions (minimum - maximum) |  | 0.001 - 73.43 | 0.001 - 11.25 | 11.26 - 49.80 | 50.10 - 73.43 | - |
| Age, years |  | 51.9 (8.0) | 51.6 (8.0) | 52.3 (8.0) | 52.9 (8.5) | **0.004** |
| Male (%) |  | 3007 (43.5) | 2065 (44.8) | 888 (40.4) | 54 (49.5) | **0.001** |
| Education** | Low (%) | 2177 (31.5) | 1297 (28.1) | 829 (37.7) | 51 (46.8) | **<0.001** |
|  | High (%) | 4510 (65.2) | 3164 (68.6) | 1291 (58.7) | 55 (50.5) |  |
|  | Missing (%) | 233 (3.4) | 152 (3.3) | 78 (3.5) | 3 (2.8) |  |
| Income | ≤ £ 30,999 (%) | 3374 (48.8) | 2286 (49.6) | 1042 (47.4) | 46 (42.2) | **0.001** |
|  | ≥ £31,000 (%) | 1929 (27.9) | 1215 (26.3) | 675 (30.7) | 39 (35.8) |  |
|  | Missing (%) | 1617 (23.4) | 1112 (24.1) | 481 (21.9) | 24 (22.0) |  |
| Current smoking (%) |  | 840 (12.2) | 454 (9.9) | 358 (16.4) | 28 (25.7) | **<0.001** |
| Body mass index, kg/m<sup>2</sup> |  | 29.5 (5.4) | 29.8 (5.3) | 28.9 (5.5) | 27.7 (4.6) | **<0.001** |
| Height, cm |  | 167 (9) | 167 (9) | 168 (9) | 168 (9) | 0.26 |
| Systolic blood pressure, mmHg |  | 140 (19) | 140 (19) | 138 (19) | 134 (18) | **<0.001** |
| Diastolic blood pressure, mmHg |  | 85 (11) | 85 (11) | 84 (11) | 82 (9) | **<0.001** |
| BP lowering medication (%) |  | 2111 (31.3) | 1495 (33.2) | 593 (27.7) | 23 (21.9) | **<0.001** |
| Type 2 Diabetes (%) |  | 331 (4.8) | 204 (4.4) | 121 (5.5) | 6 (5.5) | 0.14 |
| Prevalent coronary artery disease (%) |  | 265 (3.8) | 186 (4.0) | 76 (3.5) | 3 (2.8) | 0.43 |
| Prevalent heart failure (%) |  | 22 (0.3) | 12 (0.3) | 10 (0.5) | 0 (0.0) | 0.34 |
| Incident AF (%) |  | 205 (3.0) | 134 (2.9) | 66 (3.0) | 5 (4.6) | 0.59 |

\*European genetic ancestry categories: Low - 0.001% - 11.25%; Medium: 11.26% - 49.80%; High: 50.10% - 73.43%.
\*\*Education: High - College or University degree; NVQ, HND, or HNC or equivalent, Other professional qualifications (e.g., nursing, teaching); Low - A levels / AS levels or equivalent; O levels / GCSEs or equivalent; CSEs or equivalent; None of the above.
Abbreviations: AF, atrial fibrillation. Data are presented as mean (standard deviation) unless otherwise stated.

**Table 2.**
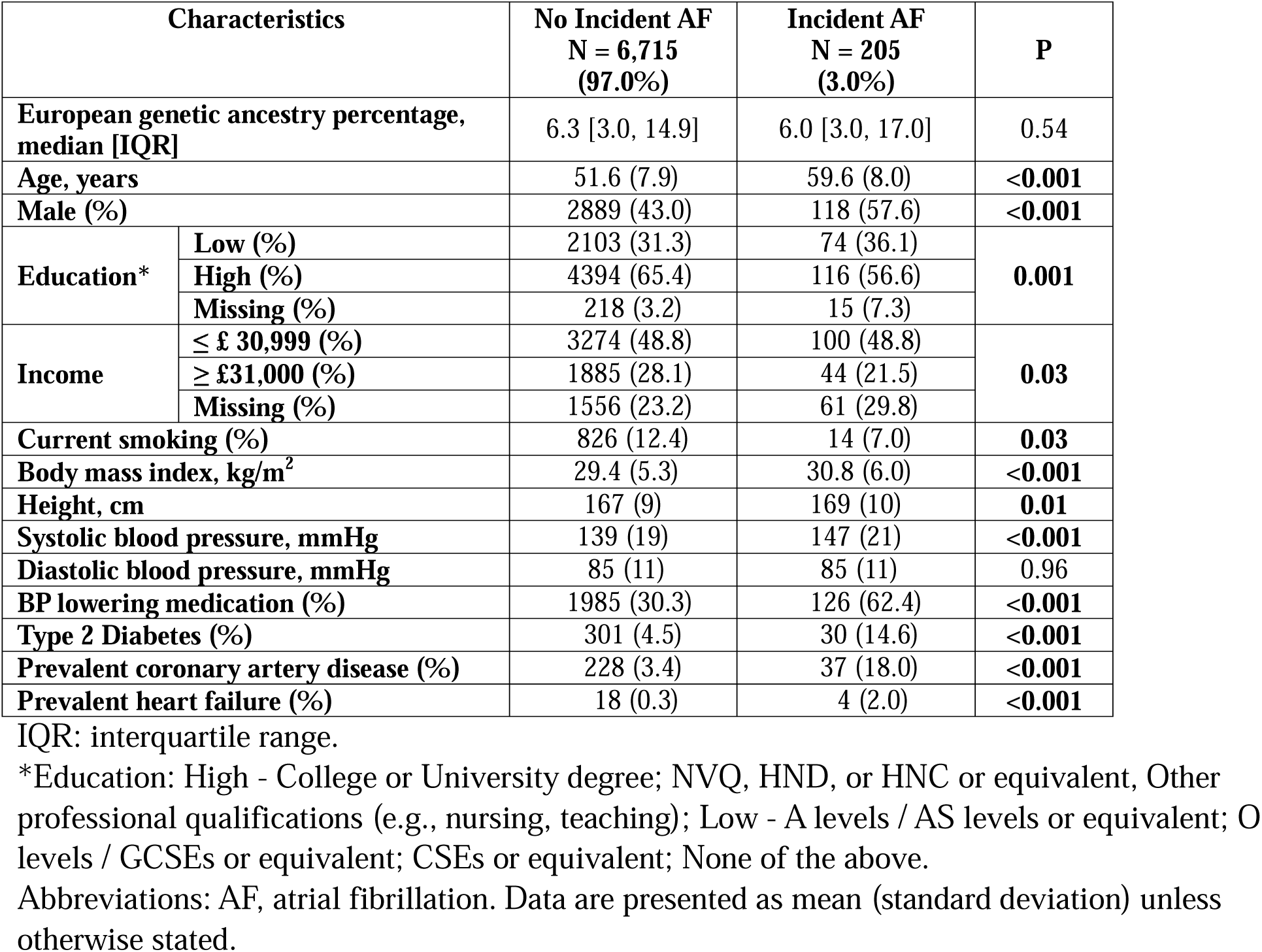
Baseline characteristics by AF incidence status.

When analyzed continuously, each 10% increase in European genetic ancestry was not significantly associated with incident AF in either the minimally adjusted Model 1 (hazard ratio [HR] 1.03, 95% confidence interval [CI]: 0.92-1.15) or the fully adjusted Model 2 (HR 1.07, 95% CI: 0.95-1.20), **Table 3**. When the European genetic ancestry was treated as a categorical variable, the incidence rate per 1,000 person-years was 2.15 in the low group, 2.23 in the middle group, and 3.36 in the high group. Compared with the low ancestry group, in Model 1, the middle group was not associated with AF risk (HR 0.99, 95% CI: 0.73-1.32), while the high ancestry group showed a nominal association (HR 1.26, 95% CI: 0.51-3.09), but the estimate was imprecise. In Model 2, both groups showed a nominal association (middle group: HR 1.06, 95% CI: 0.78-1.44; high group: HR 1.64, 95% CI: 0.66-4.04). Similar findings were observed in the sex-stratified analyses, **Supplementary Table 1**. In Model 2, the association between each 10% increase in European genetic ancestry and incident AF was nominal and not statistically significant in males (HR: 1.04, 95% CI: 0.89-1.22), females (HR: 1.12, 95% CI: 0.93-1.34) and postmenopausal females (HR 1.14, 95% CI: 0.92-1.41). When European genetic ancestry was treated as a categorical variable, the high ancestry group had slightly elevated hazard ratios in the middle and high groups, with 95% CIs covering the null. The data were sparse in both males and females with high European genetic ancestry.

**Table 3.** Association of European genetic ancestry with AF risk.

| Estimates |  | Per 10% increase in European genetic ancestry percentage | Low* | Middle* | High* |
| --- | --- | --- | --- | --- | --- |
| Model 1 | N | 6920 | 4613 | 2198 | 109 |
|  | Number of AF | 205 | 134 | 66 | 5 |
|  | Person-years | 93391 | 62334 | 29567 | 1490 |
|  | Incidence Rate per 1000 person-years | 2.20 | 2.15 | 2.23 | 3.36 |
|  | HR (95% CI) | 1.03 (0.92 - 1.15)<br>p = 0.66 | Ref | 0.99 (0.73 - 1.32)<br>p = 0.92 | 1.26 (0.51 - 3.09)<br>p = 0.61 |
| Model 2 | N | 6491 | 4320 | 2071 | 100 |
|  | Number of AF | 196 | 128 | 63 | 5 |
|  | Person-years | 87578 | 58363 | 27858 | 1357 |
|  | Incidence Rate per 1000 person-years | 2.24 | 2.19 | 2.26 | 3.68 |
|  | HR (95% CI) | 1.07 (0.95 - 1.2)<br>p = 0.27 | Ref | 1.06 (0.78 - 1.44)<br>p = 0.72 | 1.64 (0.66 - 4.04)<br>p = 0.28 |

\*European genetic ancestry categories: Low - 0.001% - 11.25%; Medium: 11.26% - 49.80%; High: 50.10% - 73.43%.
Abbreviations: AF, atrial fibrillation; HR, hazard ratio; CI, confidence interval.

The potential non-linear association between European ancestry and AF risk is shown in **Figure 2**. The HR curves showed no strong evidence of a monotonic trend, with the HRs remaining close to the null across most of the ancestry distribution. However, there was a slight elevation in risk at higher European genetic ancestry proportions. The confidence intervals were wide and crossed the null at the upper end of the distribution due to the sparse data. Sex-stratified non-linear curves showed a similar pattern, **Supplementary Figure 3**.

**Figure 2.**
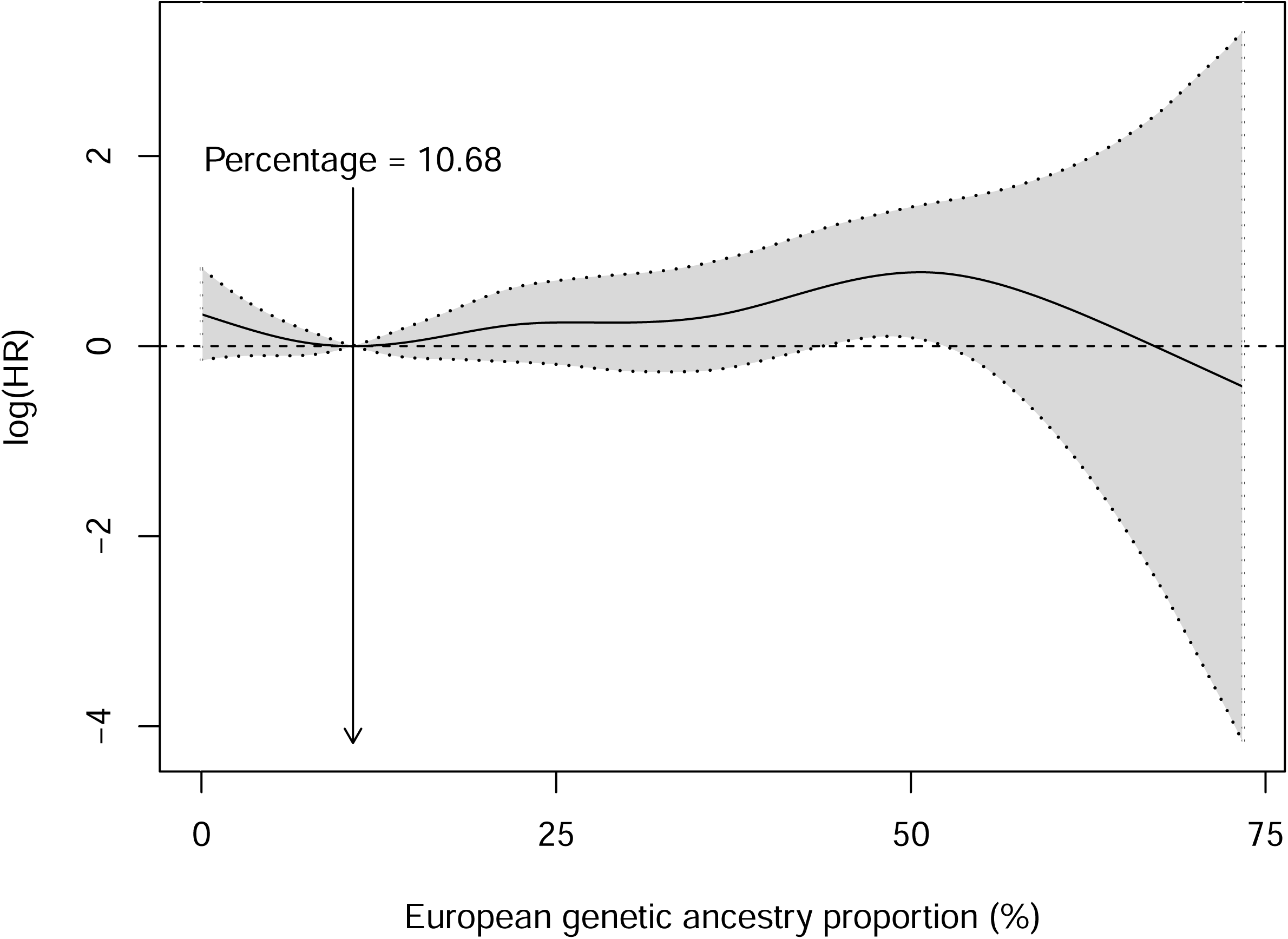
Flexible non-linear hazard ratio curves of the association between European genetic ancestry and incident AF among the UK Biobank participants self-reported as Black. Models adjusted for age at enrollment, sex (male vs. female), education (high [college or university degree; NVQ, HND, or HNC or equivalent, other professional qualifications] vs. low [ A levels / AS levels or equivalent, O levels / GCSEs or equivalent, CSEs or equivalent, none of the above] vs. missing), income (≤ £ 30,999 vs. ≥ £31,000 vs. missing), current smoking, body mass index, height, systolic blood pressure, diastolic blood pressure, blood pressure lowering medication use, prevalent type 2 diabetes, prevalent coronary artery disease, prevalent heart failure.

**Supplementary Table 2** includes general characteristics of the four meta-analyzed studies. The fixed-effect meta-analysis of all four cohorts yielded relative risk (RR) 1.03 (95 % CI: 0.99-1.08) with substantial heterogeneity (I^2^ = 72 %, p for heterogeneity = 0.01), **Figure 3A**. The random-effects summary was RR 1.09 (95 % CI: 0.99-1.21), **Figure 3B**. The leave-one-out analysis confirmed that exclusion of the WHI study materially affected both the summary estimate and between-study heterogeneity, **Supplementary Figure 4**. When WHI was removed, the fixed-effect RR increased to 1.14 (95 % CI: 1.05-1.23) and I^2^ decreased to 0 % (p for heterogeneity = 0.40), indicating homogeneity across the remaining three cohorts. In contrast, omitting any of the other studies produced minimal changes in the pooled estimate and did not materially reduce heterogeneity, suggesting that WHI was the primary source of inconsistency in the meta-analysis.

**Figure 3.**
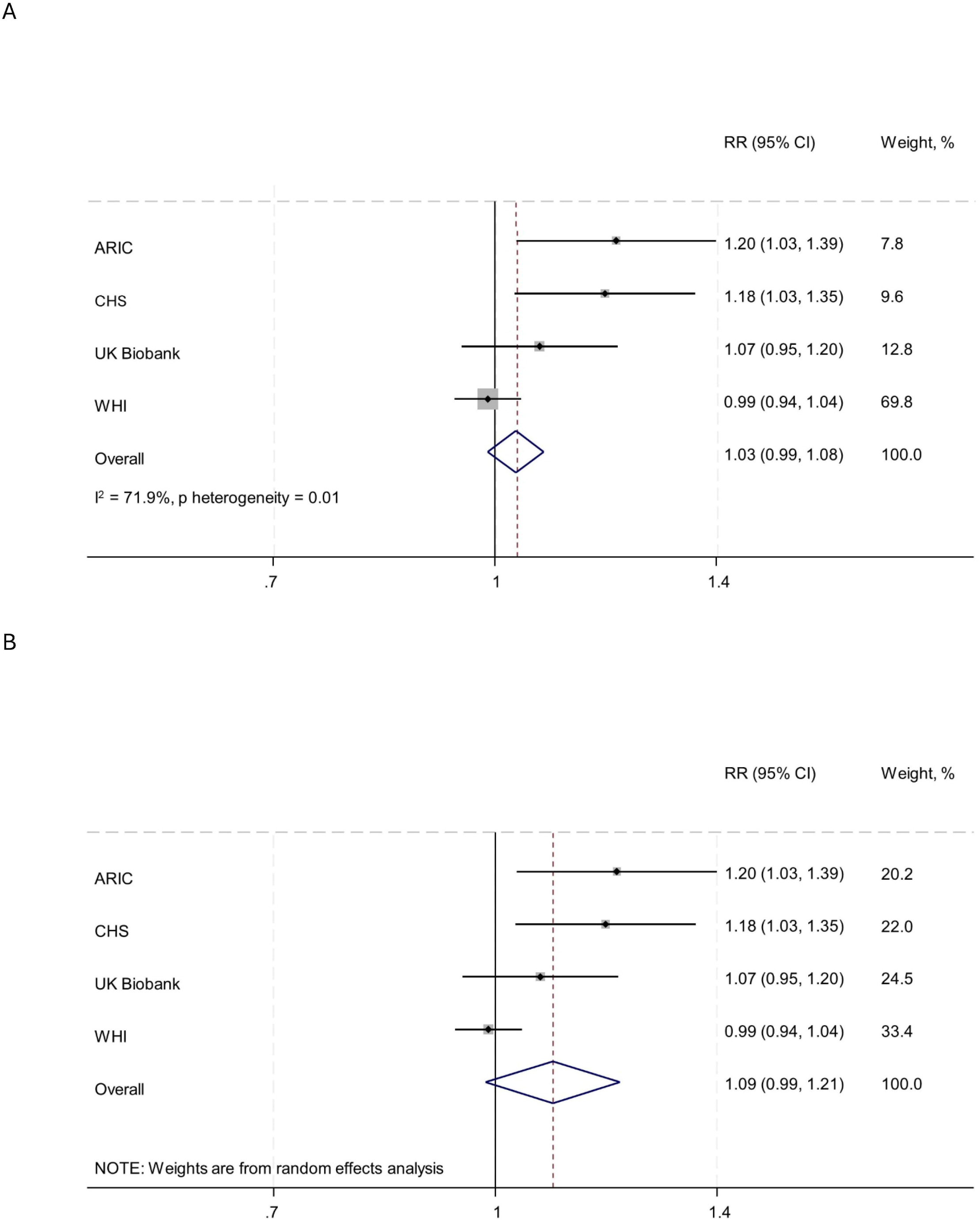
Association between genome-wide European ancestry and atrial fibrillation risk among Black adults across four cohorts. Panel A shows the fixed-effect meta-analysis; Panel B shows the random-effects meta-analysis. Relative risks (RRs) are per 10% increase in European ancestry.

## DISCUSSION

In this large, prospective analysis of UK Biobank participants who self-identified as Black, we found a suggestive association between higher European genetic ancestry and increased risk of incident AF. Individuals in the highest European ancestry group had a higher incidence rate of AF compared with those with lower ancestry proportions, but the estimates were imprecise. Our results are consistent with earlier studies from ARIC and CHS, which reported associations between European ancestry and AF among Black individuals. However, they contrast with the findings from the WHI cohort, which did not observe a similar relationship. Notably, our meta-analysis incorporating data from all four cohorts yielded a modest but non-significant pooled estimate, with substantial heterogeneity largely attributable to the WHI study. When WHI was excluded, the association between European ancestry and AF was statistically significant and heterogeneity was reduced, suggesting more consistent findings across ARIC, CHS, and UKB. These patterns support the hypothesis that European ancestry may confer increased genetic susceptibility to AF, though the magnitude of effect appears modest and may not fully explain the difference.

The observed heterogeneity in the association between European genetic ancestry and AF across cohorts, particularly between WHI^19^ and the other studies (ARIC^20^, CHS^21^, and UKB^10^), may reflect key differences in study design, population characteristics, and AF ascertainment. WHI included only postmenopausal women, whereas ARIC, CHS, and UKB included both men and women, potentially introducing sex-related biological differences in AF risk.^22^ However, the sex-stratified analysis and analysis among only postmenopausal women in UKB did not reveal meaningful differences, which may point to differences in other study-specific factors. For example, in WHI, AF was identified using hospitalization records adjudicated by the study and diagnosis codes obtained from linked Medicare claims data,^16^ which likely failed to capture asymptomatic or subclinical AF cases. In contrast, in ARIC and CHS, AF was ascertained through electrocardiograms conducted at study visits, as well as from hospitalization records and death certificates.^23–25^ In the UKB, AF was ascertained by integrating data from HES, primary care records, death registries, and self-reported medical conditions.^10^ Additionally, WHI included both prevalent and incident AF cases, while the other studies focused exclusively on incident AF, which could introduce survival or selection bias. These methodological and demographic differences across cohorts may have contributed to the variability in observed associations between European ancestry and AF risk.

It has been established that the genetic architecture of AF differs between African and European ancestry populations, and the allele frequencies at AF-associated loci may vary across these groups.^26, 27^ Additional factors may help explain the observed paradox in AF incidence across racial groups. AF is often asymptomatic and can remain undetected without continuous or systematic monitoring. Prior studies have reported that AF is underdiagnosed in Black individuals compared to White individuals.^25, 28^ Underascertainment in Black may be driven in part by reduced access to medical care and differences in healthcare utilization, all of which could contribute to the divergence in reported AF incidence.^25, 28–30^ Further, Black individuals often have a greater burden of comorbid conditions such as hypertension, diabetes, and heart failure, which are associated with increased risk of premature cardiovascular morbidity and mortality.^31, 32^ These conditions can lead to earlier occurrence of adverse events or death from causes other than AF, effectively shortening the period during which AF could be clinically detected.

Our analysis has several key strengths, including a large sample of self-identified Black individuals in the UKB with comprehensive genotype data, robust AF ascertainment through multiple linked data sources, and the capacity to model genetic ancestry both as a continuous and categorical variable. The use of spline modeling allowed for flexible detection of non-linear associations, and the meta-analysis approach enabled integration of data across multiple large cohorts to enhance generalizability. However, several limitations should be acknowledged. First, despite the relatively large overall sample, the number of AF events among individuals with high European ancestry was modest, contributing to wide confidence intervals and reduced precision. Second, heterogeneity in AF case definitions, population characteristics, and ascertainment methods across the included cohorts may have introduced variability that influenced the pooled estimates.

## CONCLUSION

In summary, our findings from the UK Biobank suggest a potential association between higher European genetic ancestry and increased AF risk among individuals self-identifying as Black. When synthesized with prior studies, the overall evidence supports a modest ancestry-related effect that may be obscured or modified by other factors in certain populations. Future studies with larger, more diverse cohorts and harmonized methodologies are needed to clarify the role of genetic ancestry in AF pathogenesis and to better understand the interplay of biological and social factors shaping racial disparities in AF risk.

## Supporting information

Supplemental results

## Data Availability

All data produced in the present study are available through the UK Biobank.

## ACKNOWLEDGEMENT

This research has been conducted using the UK Biobank Resource under Application Number 34031.

