## Supplemental results for "Genetic Ancestry and Risk of Atrial Fibrillation in Individuals of Black Ethnicity in the UK Biobank"

**Supplementary Table 1.** Sex-stratified association of European genetic ancestry with AF risk.

| **Estimates** | | | **Male** | | | | **Female** | | | | **Postmenopausal Female** | | | |
| --- | --- | --- | --- | --- | --- | --- | --- | --- | --- | --- | --- | --- | --- | --- |
|  |  |  | **Per 10% increase in European genetic ancestry percentage** | **Low*** | **Middle*** | **High*** | **Per 10% increase in European genetic ancestry percentage** | **Low*** | **Middle*** | **High*** | **Per 10% increase in European genetic ancestry percentage** | **Low*** | **Middle*** | **High*** |
| **Model 1** | **N** | 3007 | | 2065 | 888 | 54 | 3913 | 2548 | 1310 | 55 | 1682 | 1122 | 535 | 25 |
|  | **Number of AF** | 118 | | 79 | 35 | 4 | 87 | 55 | 31 | 1 | 58 | 38 | 19 | 1 |
|  | **Person-years** | 40290 | | 27694 | 11872 | 724 | 53101 | 34641 | 17695 | 766 | 22571 | 15113 | 7116 | 343 |
|  | **Incidence Rate** **per 1000 person-years** | 2.93 | | 2.85 | 2.95 | 5.53 | 1.64 | 1.59 | 1.75 | 1.31 | 2.57 | 2.51 | 2.67 | 2.91 |
|  | **HR (95% CI)** | 1.02 (0.88 - 1.18)  p = 0.8161 | | Ref | 0.95 (0.64 - 1.42)  p = 0.8136 | 1.63 (0.59 - 4.48)  p = 0.3457 | 1.05 (0.88 - 1.26)  p = 0.5924 | Ref | 1.05 (0.68 - 1.64)  p = 0.8139 | 0.71 (0.1 - 5.15)  p = 0.7358 | 1.06 (0.86 - 1.31)  p = 0.5618 | Ref | 1 (0.58 - 1.75)  p = 0.9888 | 0.91 (0.12 - 6.68)  p = 0.9282 |
| **Model 2** | **N** | 2799 | | 1925 | 826 | 48 | 3962 | 2395 | 1245 | 52 | 1615 | 1073 | 518 | 24 |
|  | **Number of AF** | 112 | | 76 | 32 | 4 | 84 | 52 | 31 | 1 | 57 | 37 | 19 | 1 |
|  | **Person-years** | 37466 | | 25794 | 11037 | 635 | 50112 | 32569 | 16822 | 722 | 21683 | 14455 | 6901 | 327 |
|  | **Incidence Rate per 1000 person-years** | 2.99 | | 2.95 | 2.90 | 6.30 | 1.68 | 1.60 | 1.84 | 1.38 | 2.63 | 2.56 | 2.75 | 3.06 |
|  | **HR (95% CI)** | 1.04 (0.89 - 1.22)  p = 0.5912 | | Ref | 1 (0.65 - 1.53)  p = 0.9943 | 1.96 (0.7 - 5.45)  p = 0.1993 | 1.12 (0.93 - 1.34)  p = 0.2369 | Ref | 1.16 (0.74 - 1.84)  p = 0.5165 | 0.85 (0.12 - 6.23)  p = 0.8727 | 1.14 (0.92 - 1.41)  p = 0.2383 | Ref | 1.11 (0.63 - 1.98)  p = 0.7122 | 1.01 (0.14 - 7.55)  p = 0.9914 |

*European genetic ancestry categories: Low - 0.001% - 11.25%; Medium: 11.26% - 49.80%; High: 50.10% - 73.43%.

Abbreviations: AF, atrial fibrillation; HR, hazard ratio; CI, confidence interval.

Model 1: adjusted for age at enrollment, education (high [college or university degree; NVQ, HND, or HNC or equivalent, other professional qualifications] vs. low [ A levels / AS levels or equivalent, O levels / GCSEs or equivalent, CSEs or equivalent, none of the above] vs. missing), income (≤ £ 30,999 vs. ≥ £31,000 vs. missing).

Model 2: adjusted for current smoking, body mass index, height, systolic blood pressure, diastolic blood pressure, blood pressure lowering medication use, prevalent type 2 diabetes, prevalent coronary artery disease, prevalent heart failure, in addition to Model 1.

**Supplementary Table 2.** Characteristics of studies included in the meta-analysis.

| **Study** | **N** | **N. AF cases** | **European Ancestry**  **(mean (SD))** | **Mean Age** | **Female %** | **Follow-up** | **RR**  **(95% CI)**  **per 10%** |
| --- | --- | --- | --- | --- | --- | --- | --- |
| ARIC | 3,481 | 181 | 17% (11%) | 53 (6) | 63% | 16 years (median) | 1.20  (1.03, 1.39) |
| CHS | 804 | 120 | 24% (15%) | 73 (6) | 62% | 10 years (median) | 1.16  (1.03, 1.31) |
| WHI | 8,119 | 558* | 17% (median) | 63 (7) | 100% | 10 years (mean) | 0.99  (0.95, 1.05) |
| UKB | 6,920 | 205 | 6% (median) | 52 (8) | 56% | 14 years (mean) | 1.07  (0.95, 1.20) |

* 423 prevalent AF, 123 incident AF.

AF: atrial fibrillation; ARIC; Atherosclerosis Risk in Communities; CHS: Cardiovascular Health Study; RR: relative risk; SD: standard deviation; UKB: UK Biobank; WHI: Women’s Health Initiative.

**Supplementary Figure 1.** Flowchart of study participants.

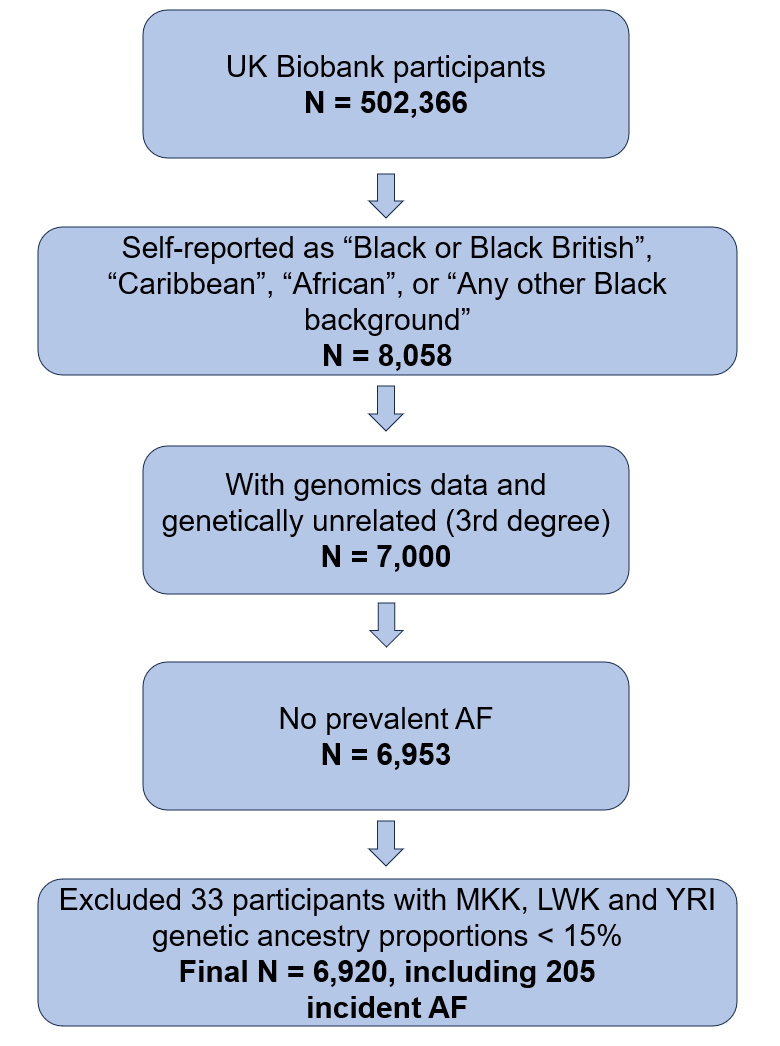

**Supplementary Figure 2.** Principal component analysis plot comparing the HapMap phase 3 reference populations and the UK Biobank self-reported Black participants. UKB: UK Biobank; CEU: Utah residents with Northern and Western European ancestry from the CEPH collection; CHB: the Han Chinese in Beijing, China; CHD: the Chinese in Metropolitan Denver, Colorado; JPT: Japanese in Tokyo, Japan; MKK: Maasai in Kinyawa, Kenya; LWK: Luhya in Webuye, Kenya; YRI: Yoruba in Ibadan, Nigeria.

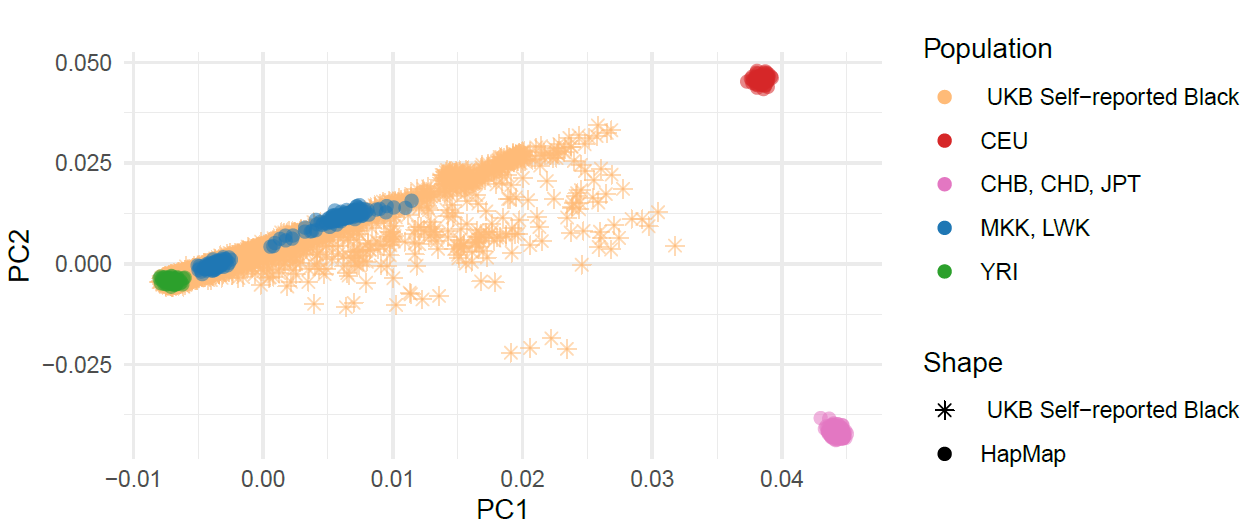

**Supplementary Figure 3.** Sex-stratified flexible non-linear hazard ratio curves of the association between European genetic ancestry and incident AF among the UK Biobank participants self-reported as Black. Models adjusted for age at enrollment, education (high [college or university degree; NVQ, HND, or HNC or equivalent, other professional qualifications] vs. low [ A levels / AS levels or equivalent, O levels / GCSEs or equivalent, CSEs or equivalent, none of the above] vs. missing), income (≤ £ 30,999 vs. ≥ £31,000 vs. missing), current smoking, body mass index, height, systolic blood pressure, diastolic blood pressure, blood pressure lowering medication use, prevalent type 2 diabetes, prevalent coronary artery disease, prevalent heart failure.

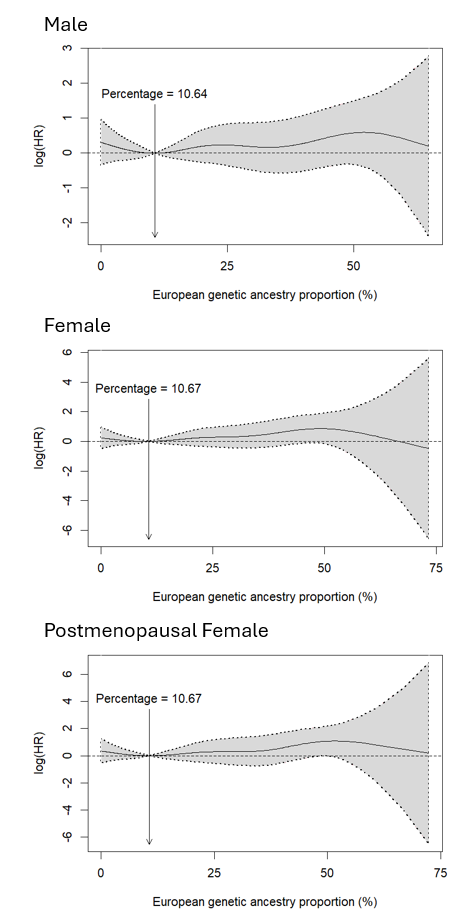

**Supplementary Figure 4.** Leave-one-out sensitivity analysis of the association between European ancestry and atrial fibrillation. Each point represents the pooled fixed-effect relative risk (RR) and 95% confidence interval after omitting one study at a time. ARIC: Atherosclerosis Risk in Communities; CHS: Cardiovascular Health Study; WHI: Women’s Health Initiative.

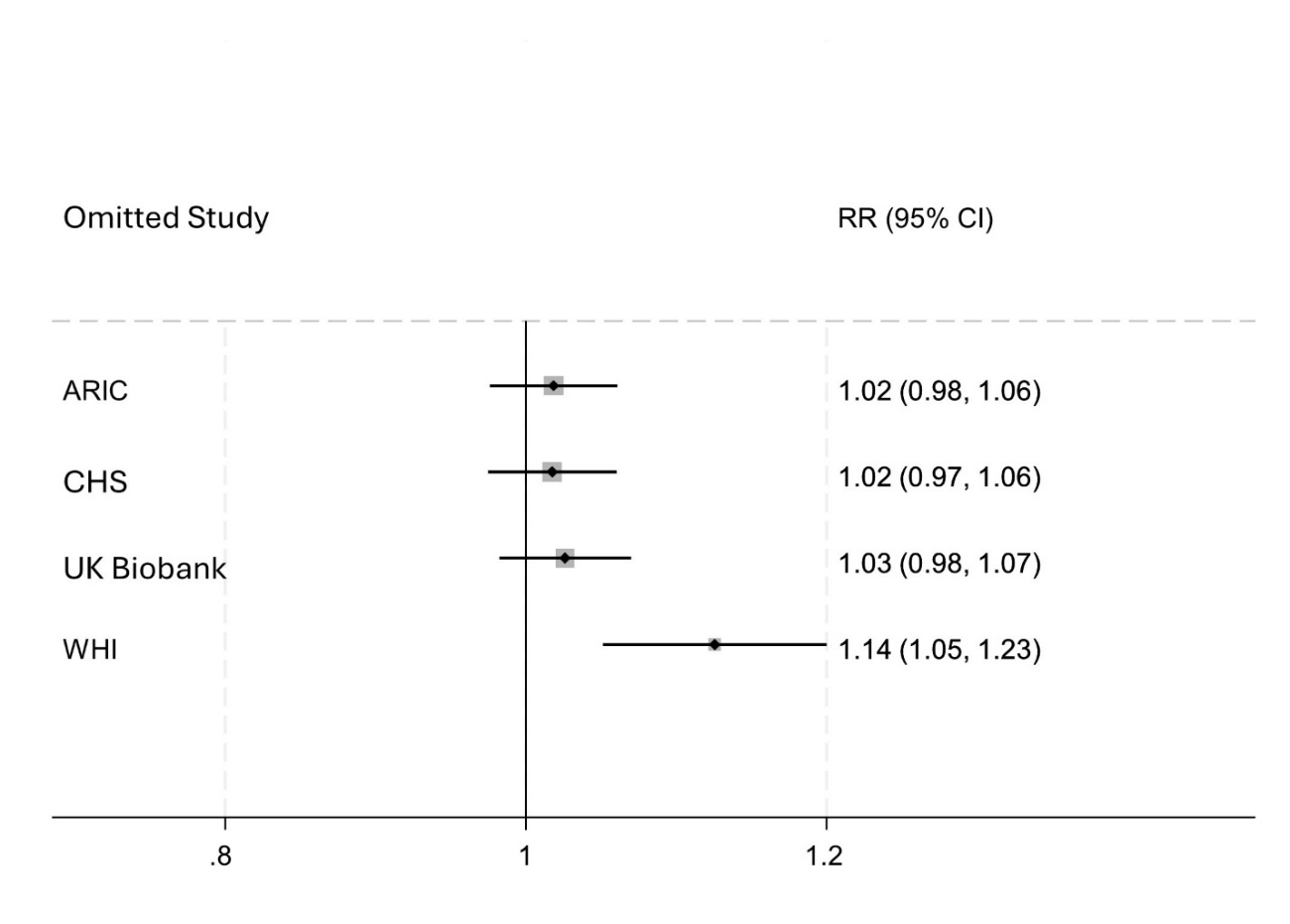
